# Prevalence and Human Health Risks of *Salmonella enterica* in Baby Poultry Sold at Agricultural Supply Stores

**DOI:** 10.64898/2026.05.04.26352231

**Authors:** K.M. Larsen, H. Blackwell, C. Patch, C. Herren, J. Bears, C.M. Armstrong, S. Kanrar, K. Harper, V. Devlin, L. Martin, O. Noyes, A. Michaelides, K. Hood, A. Lunna, A. Penny, Sarah C. Nguyen, A. J. Etter

## Abstract

Across the United States, backyard poultry (BYP) are becoming increasingly popular as a food source as well as pets. Unfortunately, they have also been a source of annual human salmonellosis outbreaks for over a decade. Previous CDC analyses suggest baby poultry are the main source of live poultry-associated outbreaks as opposed to adult birds. However, there are few data on the frequency of pathogens, such as *Salmonella enterica*, in baby poultry sold to the BYP market. Further, there is a lack of data on the serovars and antimicrobial resistance (AMR) rates in these baby poultry. We collected 643 soiled bedding and shipping box samples from agricultural supply stores primarily located in Vermont. *S. enterica* was detected in 23.5% (151/643) of samples, with the highest rates of detection in 2021-2022. Rates of *S. enterica* varied by species. Turkey poult bedding samples had the highest rates of *S. enterica* (44.4%; 8/18), while laying chick bedding samples had the lowest (19.4%; 68/350). Meat chick bedding samples had an intermediate rate, at 36% (32/89). The most common serovar detected was *Salmonella* Enteritidis, which represented 51.2% (64/125) of sequenced isolates. AMR genes or AMR-associated point mutations were detected in 21.6% (27/125) of samples, but only in non-Enteritidis serovars. These data indicate that baby poultry intended for the BYP market pose a substantial risk of salmonellosis to consumers.

## Introduction

Each year, non-typhoidal *Salmonella enterica* causes an estimated 1.3 million cases of salmonellosis, 238 deaths, and 12,500 hospitalizations in the U.S. (Scallan Walter et al., 2025). *S. enterica* is the most expensive U.S. foodborne pathogen due to its prevalence and higher hospitalization rate (Hoffmann et al., 2024; Scallan Walter et al., 2025). Estimates suggest nontyphoidal *Salmonella* costs the U.S. $17.1 billion annually, or 23% of the total costs of foodborne illness (Hoffmann et al., 2024). While most cases of salmonellosis are from contaminated food, about 11% of cases are linked to live animal contact (Hale et al., 2012), particularly contact with poultry.

According to a recent study, illnesses associated with BYP-linked outbreaks in 2015-2022 tripled compared to data from the 1990-2014 period (Basler et al., 2016; Stapleton et al., 2024). Part of this increase in illnesses was due to the steep rise in BYP purchases during the COVID-19 lockdowns, as after 2020, BYP-associated illness rates dropped slightly (Nichols et al., 2021; Stapleton et al., 2024). However, from 2021-2022, salmonellosis linked to backyard flocks across the country caused nearly 2400 confirmed illnesses (Stapleton et al., 2024). An estimated 1/29 cases of *S. enterica* infections are diagnosed and reported to public health departments, making the true illness burden in these outbreaks much higher (Scallan et al., 2011).

Despite the rise in live poultry-associated illnesses, *S. enterica* is infrequently found in adult backyard poultry. U.S. studies have found *S. enterica* in 1.9-19% of flocks (Brochu et al., 2019; Clothier, Kim, Mete, & Hill, 2018; Larsen, DeCicco, Hood, & Etter, 2022; McDonagh et al., 2019; Parzygnat et al., 2024; Patch et al., 2025; Shah et al., 2020), with most studies finding *S. enterica* in < 5% of flocks. However, a CDC review of live poultry-associated salmonellosis cases from the 1990s-2014 found that 62% of case patients reported contact with chicks or ducklings, rather than adult birds (Basler et al., 2016). Most baby poultry are purchased from agricultural feed stores, since consumers can purchase small numbers of birds more easily through stores than directly from hatcheries (Basler et al., 2016; Nichols et al., 2018). Suppliers for these stores are typically hatcheries that focus exclusively on the BYP market, which are subject to different regulations than hatcheries supplying commercial agriculture (Anonymous, 1971; Basler et al., 2016). In a 3-year study similar to ours, the Michigan Department of Health detected *S. enterica* in 35/136 (25.7%) baby poultry shipping box samples from agricultural supply stores (Sidge et al., 2019).

Due to the higher rate of salmonellosis traceable to baby poultry versus adult birds (Sidge et al., 2019), the prevalence of agricultural supply stores selling baby poultry in Vermont, and nearly 25% of Vermonters raising BYP (Larsen et al., 2022), we conducted a multi-year sampling project at Vermont agricultural supply stores to determine the prevalence of *S. enterica* in baby poultry sold to backyard poultry owners in Vermont and the serovars and antimicrobial resistance genes and mutations present.

## Methods

### *Salmonella* detection: shipping box sampling

When possible, straw shipping pads were collected directly from the empty shipping boxes at the agricultural supply stores in resealable gallon-sized freezer bags. When shipping pads were not available, a quart-sized sample of soiled bedding (straw, wood chip, paper, newspaper, or cardboard), was collected at the store, hatchery or owners’ home by our team, and placed in a clean resealable freezer bag or container. In 2020, several samples were collected by BYP owners per our instructions and left on the porch for contact-free pickup. The samples were brought back to the laboratory and frozen at −20°C or −80°C if it could not be processed immediately. The sample processing methods were based off USDA-APHIS/NPIP sampling methods (USDA-APHIS, 2010). To detect *S. enterica*, a 25-gram sample was aseptically weighed into a stomacher bag. Buffered Peptone Water (100 mL; BD Difco, Franklin Lakes, NJ) was added, and the sample was mixed in a stomacher (Seward, West Sussex, UK) for 1 minute on the standard stomaching speed or hand massaged for 2 minutes. Samples were pre-enriched at 37°C for four hours to allow bacterial recovery before proceeding with the *Salmonella* detection protocol outlined previously (Larsen et al., 2022). Briefly, 1mL of the sample was transferred into a tube containing 10mL of BD Difco™ Dehydrated Culture Media: TT (Tetrathionate) Broth Base, Hajna 500g (BD Difco, Franklin Lakes, NJ), and 100uL was transferred into a tube containing 10mL of BD Difco™ Dehydrated Culture Media: Rappaport Vassiliadis R10 (RV) Broth (BD Difco, Franklin Lakes, NJ) (Larsen et al., 2022). The RV and TT enrichments were incubated for 24 hours (Larsen et al., 2022), and the enrichments struck onto separate Xylose Lysine Tergitol-4 (XLT4) plates or *Salmonella* Non-Typhoidal Chromogenic plating media (SCPM; R&F Laboratories, Downer’s Grove, IL), incubated at 37°C (XLT-4) or 35°C (SCPM) for 24 hours, and observed for suspect *S. enterica* colonies (Larsen et al., 2022).

### *Salmonella* Confirmation

Up to four presumed positive colonies per sample were saved for PCR confirmation and further characterization. Isolates were individually grown in Tryptic Soy Broth overnight, mixed 50/50 with 50% glycerol, and stored at −80°C (Larsen et al., 2022). For *Salmonella* confirmation, suspect positive colonies were re-struck for isolation, and identified using colony pick PCR for the *hilA* gene as previously described (Larsen et al., 2022)

### Whole genome sequencing and bioinformatics analysis

Frozen isolates were transported on ice to the Vermont Department of Health Laboratory in Colchester, Vermont, where DNA extraction, library preparation, and short read whole genome sequencing (WGS) via Illumina MiSeq (Illumina, Inc., San Diego, CA) was performed as previously described (Patch et al., 2025). Seven isolates in this analysis were overnight shipped on dry ice to the USDA Agricultural Research Station (USDA-ARS) in Wyndmoor, Pennsylvania, where DNA extraction, library preparation, and amplification free long read WGS via Oxford Nanopore Technologies GridION (Oxford, United Kingdom) was performed as previously described in Patch et al. (Patch et al., 2025).

### Animal and human ethics statement

Due to the fact we sampled bedding rather than live poultry, Institutional Animal Care and Use Committee approval was not needed; Institutional Research Board approval was not required.

## Results and Discussion

### Prevalence of *S. enterica* in the Vermont Baby Poultry Market

We collected 643 shipping pad or bedding samples from 15 stores and directly sampled bedding from six local hatcheries between 2020-2023. We also sampled bedding or received bedding samples from several BYP owners with baby poultry. Bedding samples came primarily from laying chicken breeds (“layer chicks”), meat chicken breeds (“meat chicks”), ducklings, goslings, and turkey poults. We also had one quail sample and four guinea hen samples. Overall, 23.5% (151/643) of bedding samples were positive for *S. enterica* from 2020-2023, but *S. enterica* prevalence varied by year. 2020 was our pilot year for the sampling project; we sampled from two stores (one in Vermont and one in Massachusetts), one local Vermont hatchery, and received bedding samples from several individuals who had either bought from agricultural supply stores or ordered directly from hatcheries. Of the 79 samples collected in 2020, 1.3% (1/79) were positive for *S. enterica.* In 2021-2023, we sampled extensively from 15 stores and 6 hatcheries across Vermont. We collected 235 samples in 2021, of which 31.9% (75) were positive. In 2022, we collected 177 samples, and 32.8% (58) samples were positive for *S. enterica*. In 2023, we collected 152 samples, of which 11.2% (17) were positive **(Table 1)**.

**Table 1:**
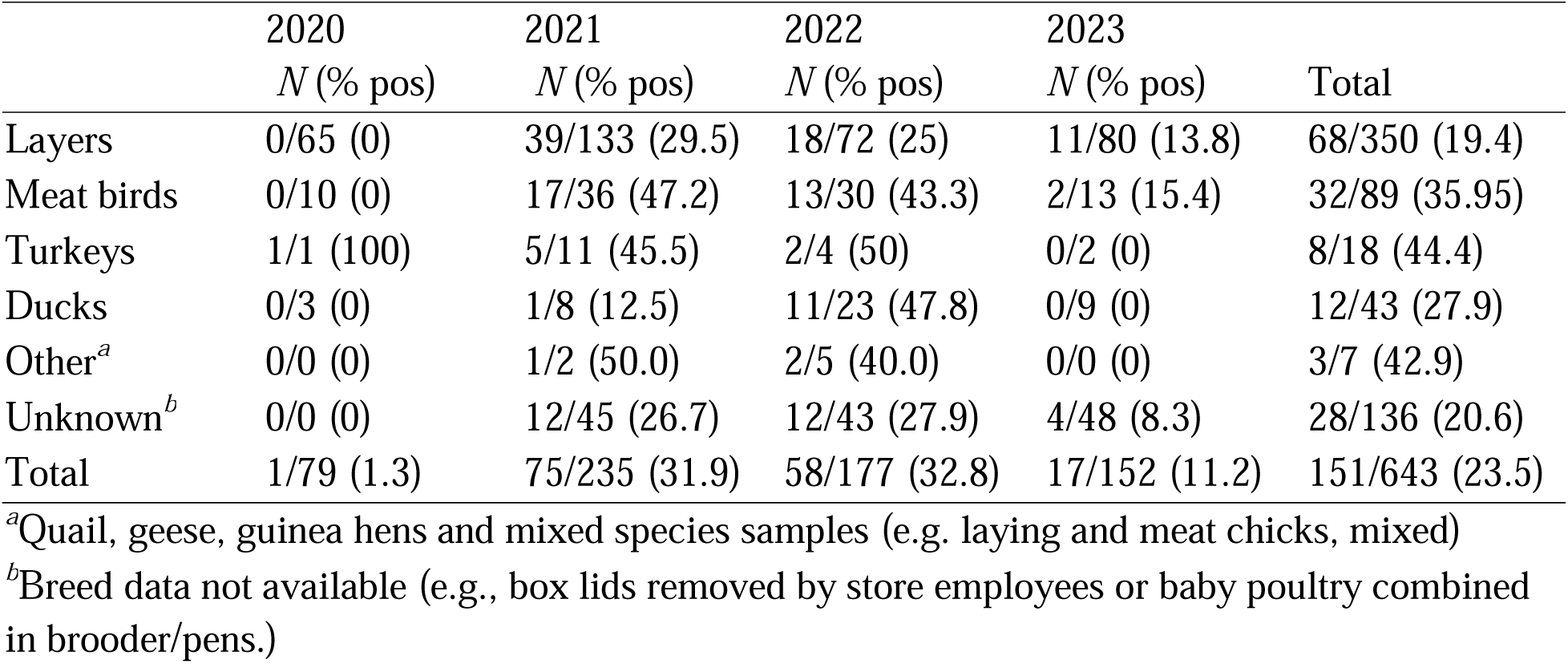
*S. enterica* prevalence in baby poultry bedding by species.

|  | 2020 | 2021 | 2022 | 2023 |  |
| --- | --- | --- | --- | --- | --- |
|  | *N* (% pos) | *N* (% pos) | *N* (% pos) | *N* (% pos) | Total |
| Layers | 0/65 (0) | 39/133 (29.5) | 18/72 (25) | 11/80 (13.8) | 68/350 (19.4) |
| Meat birds | 0/10 (0) | 17/36 (47.2) | 13/30 (43.3) | 2/13 (15.4) | 32/89 (35.95) |
| Turkeys | 1/1 (100) | 5/11 (45.5) | 2/4 (50) | 0/2 (0) | 8/18 (44.4) |
| Ducks | 0/3 (0) | 1/8 (12.5) | 11/23 (47.8) | 0/9 (0) | 12/43 (27.9) |
| Other<sup>a</sup> | 0/0 (0) | 1/2 (50.0) | 2/5 (40.0) | 0/0 (0) | 3/7 (42.9) |
| Unknown<sup>b</sup> | 0/0 (0) | 12/45 (26.7) | 12/43 (27.9) | 4/48 (8.3) | 28/136 (20.6) |
| Total | 1/79 (1.3) | 75/235 (31.9) | 58/177 (32.8) | 17/152 (11.2) | 151/643 (23.5) |

<sup>a</sup>Quail, geese, guinea hens and mixed species samples (e.g. laying and meat chicks, mixed)
<sup>b</sup>Breed data not available (e.g., box lids removed by store employees or baby poultry combined in brooder/pens.)

There are limited data on *S. enterica* prevalence in baby poultry sold to BYP market. Sidge et al. performed the only prior study of *S. enterica* surveillance using chick shipping boxes at agricultural supply stores (Sidge et al., 2019). They found an average 26% prevalence of *S. enterica* over the three-year sampling period of 2016-2018 (Sidge et al., 2019), which is slightly higher than we found. Wilkins et al. conducted an outbreak traceback investigation on a single hatchery and found no *S. enterica* in 12 cloacal samples of baby poultry at the hatchery, despite the presence of the outbreak strain in the breeder flock, hatchery, and incubator areas (Wilkins et al., 2002). Another traceback investigation, conducted in 2006, found *S. enterica* in 25/137 (18.2%) of cloacal swabs of baby poultry at agricultural supply stores in Oregon (Bidol et al., 2007).

*S. enterica* rates varied by poultry type between bedding samples from layer chicks, meat chicks, turkey poults, and ducklings. Among samples with breed/species information, bedding from layer chicks had the lowest rate (19.4%; 68/350), followed by ducklings (27.9%; 12/43), meat chicks (35.95%; 32/89), and turkey poults (44.4%; 8/18). We had only four bedding samples from guinea hen and two were positive for *S. enterica.* Our detection rate for *S. enterica* in layer chick bedding samples was only slightly higher than the rate of *S. enterica* detected in commercial laying flocks in Iowa and Pennsylvania, with detection in small flocks (<3,000 birds) at 16.7% and in medium flocks (3,000-50,000 birds) at 16% (Denagamage et al., 2016).

We found that 35.95% of meat chick bedding samples (32/89) were positive for *S. enterica*. A study of a single hatchery in Georgia (USA) supplying commercial broilers to the meat industry found that 50% of samples from post-hatching and transport carried *S. enterica* (Rothrock et al., 2024). Two other GA studies examining broiler breeder hatcheries found *S. enterica* on 12.0% (15/125) and 74.3% (26/35) of paper shipping pad samples (Cox et al., 1990; Cox, Bailey, Mauldin, Blankenship, & Wilson, 1991). A southeastern U.S. study comparing BYP flocks of broilers with commercial broiler farms found an overall prevalence rate of 19.0% for backyard farms and 52.3% for the commercial broiler farms (Parzygnat et al., 2025).

We had only 18 bedding samples from turkey poults; however, 44.4% were positive (8/18) for *S. enterica*. A higher prevalence of *S. enterica* in turkey poults than broiler chicks is the opposite of what the literature suggests for prevalence on commercial farms and at slaughter (Foley, Lynne, & Nayak, 2008; FSIS, 2021; Rostagno, Wesley, Trampel, & Hurd, 2006).

Additionally, a study in Egypt of imported and domestic turkey poults via cloacal swab found *S. enterica* in just 4% of birds sampled (Osman, Marouf, Erfan, & AlAtfeehy, 2014). Finally, a 1992 study of *S. enterica* in shipping box liners for turkey poults found just 4/22 (18.2%) were positive for *S. enterica* (Hoover, Kenney, Amick, & Hypes, 1997). It is unclear why we found higher rates of *S. enterica* than previous research.

We had 43 samples from duckling bedding, of which 12 (27.9%) were positive. There are limited studies on *S. enterica* prevalence in ducklings. A study of Belgian mule ducks raised for *foie gras* found that 50% of duckling shipment samples from French hatcheries carried *S. enterica* at arrival on rearing farms; however, by the end of rearing, only 3% were positive for *S. enterica* (Flament, Soubbotina, Mainil, & Marlier, 2012). A study in Taiwan found *S. enterica* in cloacal swabs of 37.5% of healthy week-old ducklings (Yu et al., 2008). Finally, a recent study in China found that 38.6% of fertilized eggs in a duck hatchery contained *S. enterica,* indicating a high likelihood of vertical transmission from the breeder flocks (Kang et al., 2022).

For a subset of samples (N=269) from 2021-2022, we were able to determine whether the source hatcheries participated in the National Poultry Improvement Plan (NPIP) *Salmonella* Monitoring Program (SMP) using NPIP registration sheets located on the NPIP website (https://www.poultryimprovement.org/statesContent.cfm) and cross-referencing with breed information on the baby poultry shipping box **(Table 2)**. Of our 269 shipping box bedding samples with SMP participation data for the hatchery and breed, 168 were from hatcheries and breeds enrolled in the SMP, while 101 were from hatcheries/breeds not enrolled in the SMP. *S. enterica* rates were slightly lower among non-SMP samples, with 32.7% (55/168) of samples from SMP-enrolled hatcheries and 29.7% (30/101) of samples from non-SMP hatcheries being positive for *S. enterica.* However, for layer chick bedding samples, the rate of *S. enterica* was much lower in SMP-enrolled hatcheries; 20.9% (19/91) of samples from SMP-enrolled hatcheries were positive for *S. enterica,* while 33.3% (23/69) of samples from non-SMP-enrolled hatcheries were positive. We did not have sufficient samples to assess other poultry types.

**Table 2:** *S. enterica* prevalence by year and hatchery NPIP *Salmonella* monitoring program enrollment status.

|  | SMP hatchery | non-SMP hatchery |
| --- | --- | --- |
|  | Positives (%) | Positives (%) |
| Layer breeds | 19/91 (20.9) | 23/69 (33.3) |
| All samples | 55/168 (32.7) | 30/101 (29.7) |

We could not investigate the source of the *S. enterica* in our samples, as most of our hatcheries were located out of state. However, numerous outbreak investigations have established hatcheries as the source of *S. enterica* contamination in baby poultry sold at agricultural supply stores (Anderson et al., 2016; Gaffga et al., 2012; Loharikar et al., 2012; Wilkins et al., 2002). Transmission dynamics within hatcheries are complicated. For example, vertical transmission of *S. enterica* through internally contaminated eggs is known to occur with *Salmonella* Enteritidis and Heidelberg (Barnhart, Dreesen, Bastien, & Pancorbo, 1991; Barnhart, Dreesen, & Burke, 1993), so if breeder flocks supplying the hatcheries have a high frequency of ovarian *S. enterica* it may be passed on to the baby poultry. However, most studies on vertical transmission in poultry indicate that it is a relatively rare occurrence (Wang et al., 2023; Withenshaw et al., 2021). A recent meta-analysis on commercial broiler hatcheries determined that across the pre-harvest broiler industry, the hatchery environment, rather than litter in grow houses, feed, feces, or drinking water, yielded the highest rates of *S. enterica,* at 48% (Wang et al., 2023). This was in agreement with a traceback investigation of a small hatchery supplying BYP flocks in 1999-2000, where the highest rates of *S. enterica* were found in the hatchery and incubator areas (Wilkins et al., 2002). Key sources of contamination in hatcheries include dust and “airborne fluff” that can easily disseminate between rooms in the hatchery and retain viable *S. enterica* for several years (Bailey, Cox, & Berrang, 1994; Miura, Sato, & Miyamae, 1964). *S. enterica* has been experimentally shown to spread between baby poultry in adjacent hatching trays in an incubator, independent of physical contact or the relative position of racks with contaminated eggs (above/below) in the hatcher, which suggests airborne transmission of *S. enterica* between baby poultry during hatching (Cason, Cox, & Bailey, 1994).

Baby poultry are typically shipped at one day old, in cardboard boxes containing about 100 chicks (25 per quarter) (Basler et al., 2016; Behravesh, Brinson, Hopkins, & Gomez, 2014). Ducklings, goslings, and turkey poults are shipped in smaller numbers per box due to their larger size (Basler et al., 2016). Boxes may contain multiple breeds in separate compartments, with transport conditions augmenting potential cross-contamination between sections (Basler et al., 2016). Baby poultry are most vulnerable to infection with *S. enterica* in the first few days of life and the stress of shipping may increase fecal shedding in infected chicks (Basler et al., 2016; Cox et al., 1990); consequently, exposure at the hatchery followed by close contact during shipment provides ideal circumstances for the horizontal transmission of *S. enterica* to occur.

Further spread of *S. enterica* between baby poultry may occur at stores, as stores that stock baby poultry for days or months often combine breeds as baby poultry are sold off. It is also unclear how thoroughly (if at all) stores clean and sanitize tubs or cages between poultry shipments, despite CDC recommendations to do so (Nichols et al., 2018), which may spread *S. enterica* between shipments. Additional studies are needed since it was outside the scope of this research to conduct environmental sampling on the cages and tubs to assess this. One of the stores in our study, which only sold baby poultry via customer pre-order, moved the chicks to stacked brooders and then gave the shipping boxes to the customers to transport the baby poultry home. This could spread *S. enterica* as well if uninfected baby poultry are put in a box with *S. enterica-*containing bedding. Overall, data suggests that while the hatcheries are the initial infection source, store practices can provide opportunities for increased cross-infection among the baby poultry.

### Serovars of *S. enterica* present in baby poultry

We sequenced 163 *S. enterica* isolates representing 127 samples (for some samples, we sequenced more than one isolate; **Table S1**) in collaboration with the Vermont Department of Health Laboratory and USDA-ARS. For samples with multiple sequenced isolates, all isolates were the same serovar and carried the same AMR genes. Consequently, we did not investigate these isolate genomes further. Of the remaining 127 unique genomes, two genomes were discarded from further analyses due to their low quality. The majority of isolates (64/125) were *Salmonella* Enteritidis, followed by *Salmonella* Infantis (18/125), Kentucky (10/125) and Alachua (9/125), with the remaining 24 isolates belonging to 10 other serovars (**Figure 1**). We did not find *Salmonella* Typhimurium in our samples, but four samples were from serovar I 4,[5],12:i-(monophasic *Salmonella* Typhimurium). Notably, *Salmonella* Alachua was primarily found in one hatchery and was only found in 2023. Its appearance in this hatchery coincided with a drop in *S. enterica* prevalence among samples from this hatchery, suggesting increased hatchery biosecurity measures possibly following a depopulation event. Overall, bedding from layer chicks had the widest variety of serovars, with *Salmonella* Enteritidis (24/53), *Salmonella* Infantis (9/53), and *Salmonella* Alachua (7/53) being the most commonly identified serovars **(Table 3)**.

**Figure 1:**
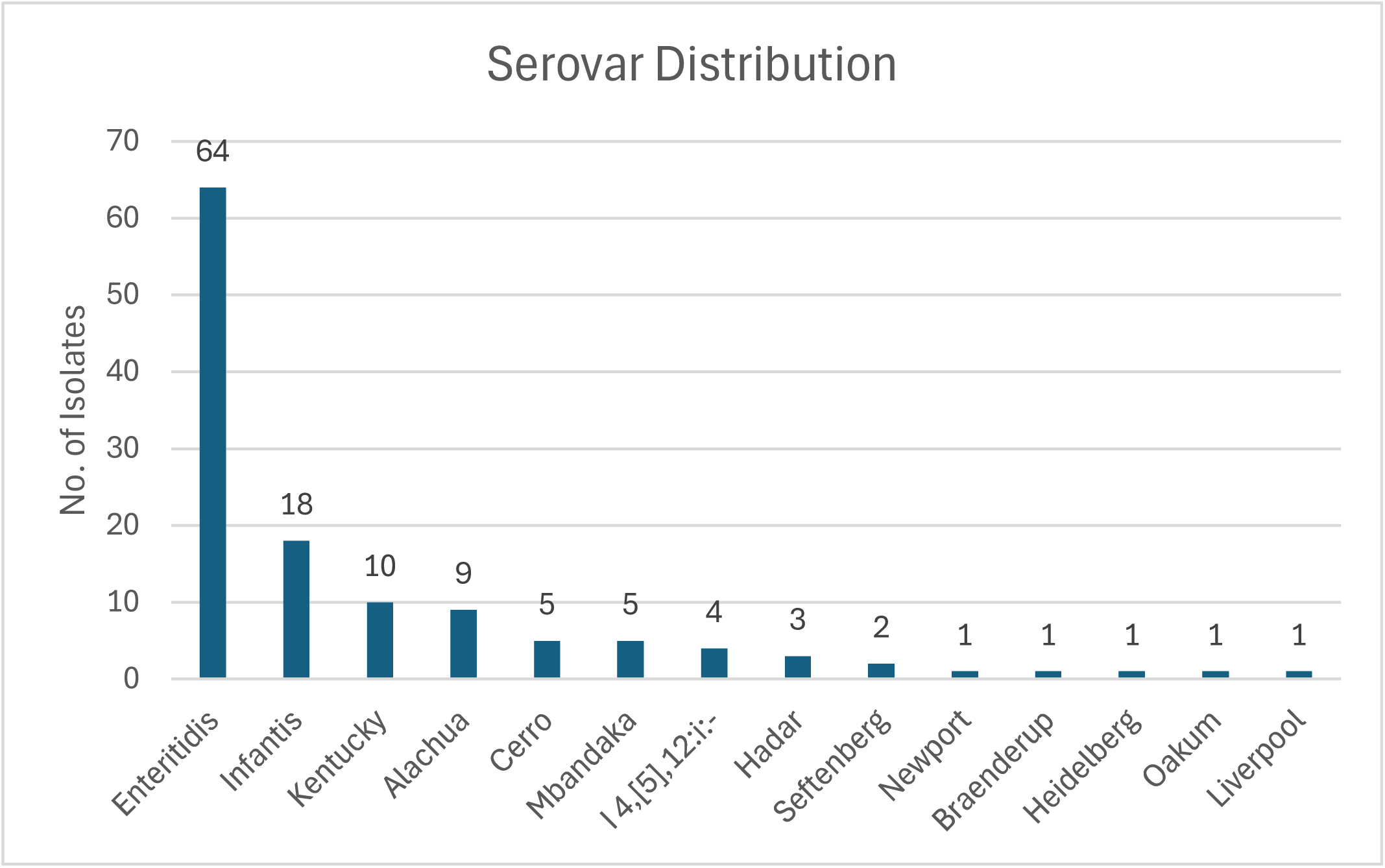
*S. enterica* serovars present in baby poultry sold in agricultural supply stores.

**Table 3:** *S. enterica* serovar distribution by species bedding samples.

|  | Layer | Meat | Chicken<br>(unknown) | Turkey | Duck | Unknown | Total |
| --- | --- | --- | --- | --- | --- | --- | --- |
| Enteritidis | 24 (45.3) | 12 (75.0) | 14 (51.9) | 0 (0) | 4 (36.4) | 10 (71.4) | 64 (51.2) |
| Cerro | 2 (3.8) | 0 (0) | 3 (11.1) | 0 (0) | 0 (0) | 0 (0) | 5 (4.0) |
| Hadar | 0 (0) | 0 (0) | 1 (3.7) | 2 (40.0) | 0 (0) | 0 (0) | 3 (2.4) |
| Heidelberg | 1 (1.9) | 0 (0) | 0 (0) | 0 (0) | 0 (0) | 0 (0) | 1 (0.8) |
| I 4,[5],12:i:- | 2 (3.8) | 0 (0) | 0 (0) | 0 (0) | 1 (9.1) | 1 (7.1) | 4 (3.2) |
| Infantis | 9 (17.0) | 0 (0) | 1 (3.7) | 2 (40.0) | 4 (36.4) | 2 (14.3) | 18 (14.4) |
| Kentucky | 4 (7.5) | 3 (18.8) | 3 (11.1) | 0 (0) | 0 (0) | 0 (0) | 10 (8.0) |
| Liverpool | 1 (1.9) | 0 (0) | 0 (0) | 0 (0) | 0 (0) | 0 (0) | 1 (0.8) |
| Mbandaka | 0 (0) | 0 (0) | 2 (7.4) | 1 (20.0) | 2 (18.2) | 0 (0) | 5 (4.0) |
| Newport | 0 (0) | 0 (0) | 0 (0) | 0 (0) | 0 (0) | 1 (7.1) | 1 (0.8) |
| Ouakam | 0 (0) | 0 (0) | 1 (3.7) | 0 (0) | 0 (0) | 0 (0) | 1 (0.8) |
| Seftenberg | 2 (3.8) | 0 (0) | 0 (0) | 0 (0) | 0 (0) | 0 (0) | 2 (1.6) |
| Alachua | 7 (13.2) | 1 (6.3) | 1 (3.7) | 0 (0) | 0 (0) | 0 (0) | 9 (7.2) |
| Braenderup | 0 (0) | 0 (0) | 1 (3.7) | 0 (0) | 0 (0) | 0 (0) | 1 (0.8) |
| Total | 52 | 16 | 27 | 5 | 11 | 14 | 125 |

For meat chick bedding samples, *Salmonella* Enteritidis (12/16) was the most common, while *Salmonella* Kentucky (3/16) and *Salmonella* Alachua (1/16) were relatively uncommon. Studies using deep serotyping have identified substantial variation in population structures within different farms and even grow houses, with some farms/houses having *Salmonella* Enteritidis as the chief serovar though a majority had *Salmonella* Kentucky as the chief serovar (Obe, Siceloff, Crowe, Scott, & Shariat, 2023). *Salmonella* Alachua is relatively rare in broilers at the slaughter level, which is similar to our finding (Siceloff, Waltman, & Shariat, 2022).

Turkey poult bedding samples carried *Salmonella* Hadar (2/5), *Salmonella* Infantis (2/5) or *Salmonella* Mbandaka (1/5). The presence of *Salmonella* Hadar aligns with historic trends in commercial turkeys (Jackson, Griffin, Cole, Walsh, & Chai, 2013), while the presence of *Salmonella* Infantis aligns with recent trends in turkeys (Ellington, Hebron, Crespo, & Machado, 2021; McMillan, Weinroth, & Frye, 2022). Notably, in commercial turkeys, *Salmonella* Hadar was considered a serovar of “public health significance” in the now-withdrawn 2024 FSIS proposed poultry standards (Anonymous, 2024). Neither *Salmonella* Typhimurium nor *Salmonella* Muenchen, the other proposed serovars of “public health significance” (Anonymous, 2024), were found in our samples.

Duckling bedding samples contained *Salmonella* Enteritidis (4/11), *Salmonella* Infantis (4/11) or *Salmonella* Mbandaka (2/11); one contained *Salmonella* I 4,[5],12:i-. This is in direct contrast to the distribution found in Chinese duckling hatcheries, where *Salmonella* Anatum and *Salmonella* Typhimurium predominated in the hatchery environment and Enteritidis, Infantis, and Mbandaka were not found (Kang et al., 2022). It also differs from the Belgian study, which found *Salmonella* Indiana and *Salmonella* Regent were most common, *Salmonella* Mbandaka and Enteritidis were rare, and *Salmonella* Infantis was absent (Flament et al., 2012). A study in Taiwan only differentiated down to serogroups and found that serogroup C1 (which contains *Salmonella* Infantis) predominated (Yu et al., 2008).

*Salmonella* Enteritidis was identified in bedding from all species except turkey poults, while *Salmonella* Infantis was found in all sample types except meat chicks. *Salmonella* Mbandaka was found in bedding samples from all except laying chicks and meat chicks.

*Salmonella* Kentucky was surprisingly rare; it was primarily found in three bedding samples from meat chicks, four from laying chicks, and three from chick bedding samples without breed information. *Salmonella* Hadar was also rare; it was found in two bedding samples from turkey poults (2/3) and one chicken sample without breed information. *Salmonella* Heidelberg, which historically caused outbreaks from both eggs and poultry meat, and is known to be vertically transmitted (Barnhart et al., 1993; CDC, 1986, 2012, 2014; Gieraltowski et al., 2016), was only found in one sample from laying chicks. *Salmonella* Kentucky is known to be highly prevalent in commercial meat birds (Jones et al., 2008); therefore, it was not surprising to find it in bedding samples from backyard poultry meat chicks, although its low frequency was unexpected.

Interestingly, we did not find *Salmonella* Infantis in bedding from meat chicks, despite its rise in commercial birds (Mattock et al., 2024; McMillan et al., 2022) and its presence in all other species sampled within this study. *Salmonella* Enteritidis has been the most common serovar detected in BYP-associated outbreaks (30.5% of infections), followed by *Salmonella* Hadar (23.7%) and *Salmonella* Infantis (14.6%) (Stapleton et al., 2024). Our samples had a slightly different distribution **(Table 4)**, with 51.2% of the baby poultry bedding positive for *S. enterica* containing *Salmonella* Enteritidis, 2.4% containing *Salmonella* Hadar, and 14.4% yielding *Salmonella* Infantis. We found a slightly higher rate of *Salmonella* Mbandaka (4.0% vs 3.3%) and I 4,[5],12:i-(3.2% vs 1.4%) than reported for humans in BYP-associated outbreaks (Stapleton et al., 2024). We also found 8.0% of our samples contained *Salmonella* Kentucky (10/125) and 4% contained *Salmonella* Cerro (5/125). Given the rare association of these serovars with human illness, it was not alarming that they were not reported in recent BYP-associated outbreaks (Jones et al., 2008; Stapleton et al., 2024).

**Table 4:**
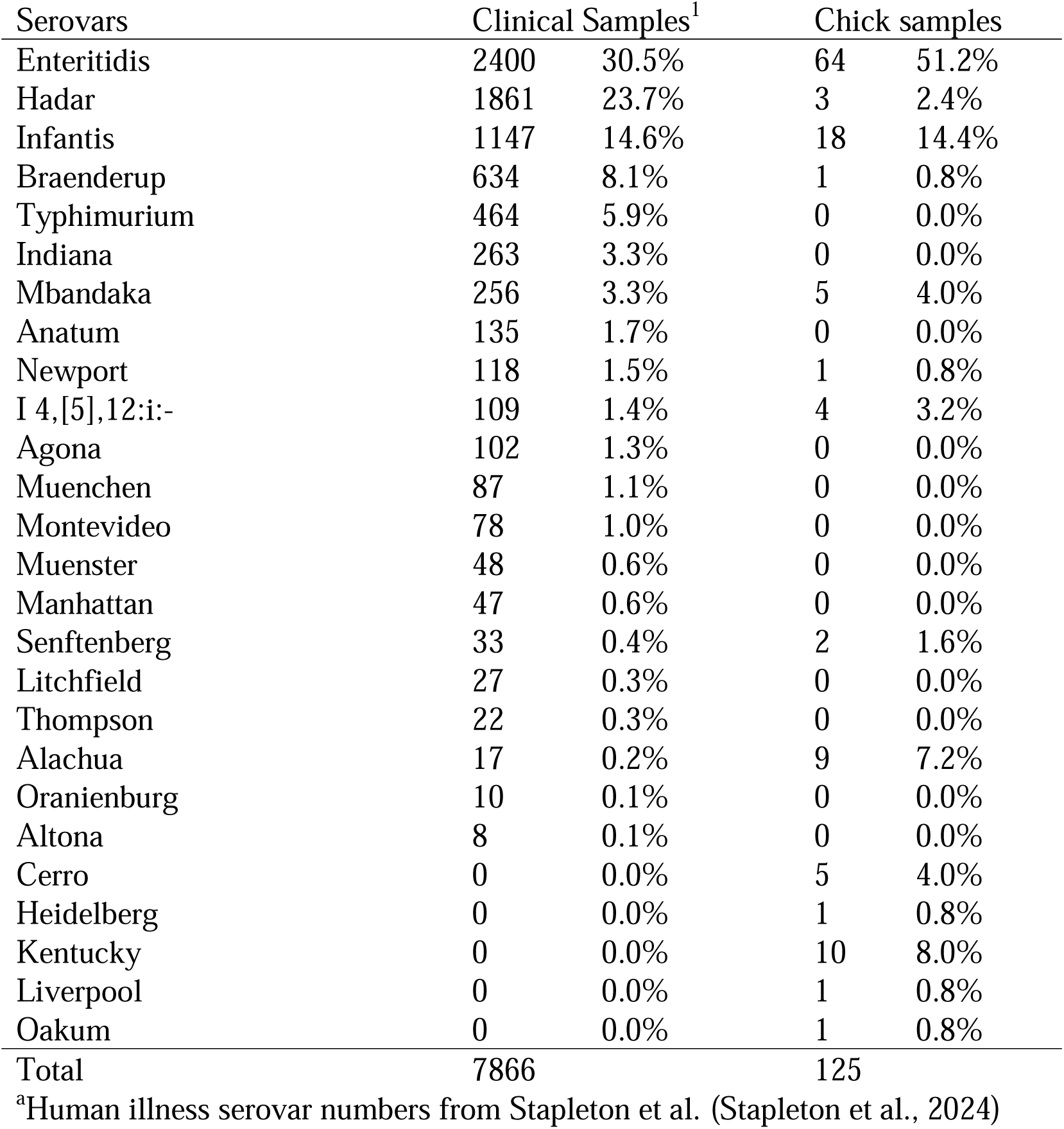
Comparison of serovar frequency in chick samples and clinical samples taken during multistate live poultry-associated outbreak investigations (Stapleton et al., 2024).

| Serovars | Clinical Samples<sup>1</sup> |  | Chick samples |  |
| --- | --- | --- | --- | --- |
| Enteritidis | 2400 | 30.5% | 64 | 51.2% |
| Hadar | 1861 | 23.7% | 3 | 2.4% |
| Infantis | 1147 | 14.6% | 18 | 14.4% |
| Braenderup | 634 | 8.1% | 1 | 0.8% |
| Typhimurium | 464 | 5.9% | 0 | 0.0% |
| Indiana | 263 | 3.3% | 0 | 0.0% |
| Mbandaka | 256 | 3.3% | 5 | 4.0% |
| Anatum | 135 | 1.7% | 0 | 0.0% |
| Newport | 118 | 1.5% | 1 | 0.8% |
| I 4,[5],12:i:- | 109 | 1.4% | 4 | 3.2% |
| Agona | 102 | 1.3% | 0 | 0.0% |
| Muenchen | 87 | 1.1% | 0 | 0.0% |
| Montevideo | 78 | 1.0% | 0 | 0.0% |
| Muenster | 48 | 0.6% | 0 | 0.0% |
| Manhattan | 47 | 0.6% | 0 | 0.0% |
| Senftenberg | 33 | 0.4% | 2 | 1.6% |
| Litchfield | 27 | 0.3% | 0 | 0.0% |
| Thompson | 22 | 0.3% | 0 | 0.0% |
| Alachua | 17 | 0.2% | 9 | 7.2% |
| Oranienburg | 10 | 0.1% | 0 | 0.0% |
| Altona | 8 | 0.1% | 0 | 0.0% |
| Cerro | 0 | 0.0% | 5 | 4.0% |
| Heidelberg | 0 | 0.0% | 1 | 0.8% |
| Kentucky | 0 | 0.0% | 10 | 8.0% |
| Liverpool | 0 | 0.0% | 1 | 0.8% |
| Oakum | 0 | 0.0% | 1 | 0.8% |
| Total | 7866 |  | 125 |  |

<sup>a</sup>Human illness serovar numbers from Stapleton et al. (Stapleton et al., 2024)

### Antimicrobial resistance (AMR) genes and point mutations

Within this study, 21.6% (27/125) of the isolates encoded at least one AMR-encoding gene. The most common AMR genes were the aminoglycoside resistance genes, *aph(6)-1d* at 20.0% (25/125) and *aph(3’’)-1b* at 17.6% (22/125), and the tetracycline resistance gene, *tetB,* at 15.2% (19/125). Fosfomycin resistance genes (*fosA7* and *fosA7.*8) were found in 10 isolates (8.0%), sulfonamide resistance genes (*sul1* or *sul2*) were found in seven isolates (5.6%), beta lactamase *bla_TEM-1_* was found in four isolates (3.2%), and fluoroquinolone resistance-encoding point mutation GyrA S83Y was found in three isolates (2.4%).

Prevalence of AMR genes varied by breed and serovar. None of the *Salmonella* Enteritidis isolates had acquired AMR genes or chromosomal point mutations. Isolates of *Salmonella* I 4,[5],12:i:-, Alachua, and Kentucky contained the most acquired AMR genes. *Salmonella* I 4,[5],12:i:-isolates carried 4-8 AMR genes, with all four isolates containing *aph(3”)-1b (strA), aph(6)-1d (strB), sul2,* and *blaTEM-1,* encoding resistance to streptomycin, kanamycin, sulfamethoxazole, ampicillin, amoxicillin, and cephalothin*. Salmonella* Alachua isolates all yielded a *strAB-fosA7.8-tetB* genotype, encoding for a streptomycin-tetracycline-kanamycin resistance phenotype. *Salmonella* Kentucky isolates mostly (7/10) carried a *strAB-tetB* genotype, while *Salmonella* Hadar all encoded a streptomycin-tetracycline-ciprofloxacin resistance phenotype (*strB-tet(A)-gyrA_S83Y* genotype). Interestingly, all *Salmonella* Infantis isolates were genetically pansusceptible.

Despite the predominance of genetically pansusceptible *Salmonella* Enteritidis, isolates that did contain AMR genes were most likely from laying hen chick bedding with 26.4% (14/53) carrying at least one AMR gene. This was closely followed by chick bedding samples without breed information (25.9%; 7/27). Despite the reported high frequency of AMR in poultry meat samples (M’Ikanatha N et al., 2010; Punchihewage-Don, Schwarz, Diria, Bowers, & Parveen, 2023), only 2/16 (12.5%) isolates from meat chick bedding carried AMR genes. Forty percent of turkey poult bedding samples (2/5) carried AMR genes and both isolates also carried a GyrA S38Y mutation conferring nalidixic acid/ciprofloxacin non-susceptibility. Duckling bedding samples had the lowest rates of AMR gene carriage at 9.1% (1/11); this isolate had a *strAB, bla_TEM-1_, tetB* genotype.

Gene carriage varied by species. Bedding samples from all species had at least one isolate carrying *aph(6’)-1d (strB)* and all species except turkey poults carried *aph(3”)-Ib (strA)* and *tetB* in at least one sample. Beta-lactamase (*bla_TEM_*) genes were found in *S. enterica* bedding samples from layer chicks, ducklings, and unknown samples **(Table 5)**, while *gyrA* point mutations were only found in *Salmonella* Hadar isolates from turkey poult bedding and one isolate from unspecified chick bedding isolates. The colistin resistance gene *mcr-9.1* was found in one *Salmonella* Liverpool isolate from laying chick bedding, however, this gene has been shown to not provide colistin resistance (Carroll et al., 2019; Macesic et al., 2021).

**Table 5:** Prevalence & classes of AMR genes in genomes of *S. enterica* in this study.

|  | Aminoglycosides |  |  |  | Beta-lactams | Colistin/<br>Polymyxin | Fluoroquinolone | Fosfomycin |  | Sulfonamides |  | Tetracyclines |  |
| --- | --- | --- | --- | --- | --- | --- | --- | --- | --- | --- | --- | --- | --- |
|  | *aph(6)-Id*<br>(*strB*) | *aph(3'')-*<br>*Ib (strA)* | *aac(3)-*<br>*VIa* | *aadA1* | *bla<sub>TEM-1</sub>* | *mcr-9.1*<sup>a</sup> | *gyrA_S83Y* | *fosA7* | *fosA7.8* | *sul1* | *sul2* | *tet(A)* | *tet(B)* |
|  | N (%) | N (%) | N (%) | N (%) | N (%) | N (%) | N (%) | N (%) | N (%) | N (%) | N (%) | N (%) | N (%) |
| Layer<br>(n=53) | 13 (24.5) | 13 (24.5) | 0 (0) | 0 (0) | 2 (3.8) | 1 (2) | 0 (0) | 1 (1.9) | 7 (13.2) | 0 (0) | 2 (3.8) | 0 (0) | 11<br>(20.8) |
| Meat<br>(n=16) | 2 (12.5) | 2 (12.5) | 0 (0) | 0 (0) | 0 (0) | 0 (0) | 0 (0) | 0 (0) | 1 (6.3) | 0 (0) | 0 (0) | 1 (6) | 2 (12.5) |
| Chicken<sup>b</sup><br>(n=27) | 6 (22.2) | 5 (18.5) | 1 (3.7) | 1 (3.7) | 0 (0) | 0 (0) | 1 (3.7) | 1 (3.7) | 0(0) | 1 (3.7) | 1 (3.7) | 2 (7.4) | 4 (14.8) |
| Turkey<br>(n=5) | 2 (40) | 0 (0) | 0 (0) | 0 (0) | 0 (0) | 0 (0) | 2 (40) | 0 (0) | 0 (0) | 0 (0) | 0 (0) | 2 (40) | 0 (0) |
| Duck<br>(n=11) | 1 (9.1) | 1 (9.1) | 0 (0) | 0 (0) | 1 (9.1) | 0 (0) | 0 (0) | 0 (0) | 0 (0) | 0 (0) | 1 (9.1) | 0 (0) | 1 (9.1) |
| Unknown<br>(n=14) | 1 (7.1) | 1 (7.1) | 1 (7.1) | 1 (7.1) | 1 (8) | 0 (0) | 0 (0) | 0 (0) | 0 (0) | 1 (7.1) | 1 (7.1) | 0 (0) | 1(7.1) |
| Total<br>(n=125) | 25 (20.0) | 22 (17.6) | 2 (1.6) | 2 (1.6) | 4 (3.2) | 1 (0.8) | 3 (2.4) | 2 (1.6) | 8 (6.3) | 2 (1.6) | 5 (4.0) | 5 (4.0) | 19<br>(15.2) |

<sup>a</sup>Does not encode colistin resistance (Carroll et al., 2019; Macesic et al., 2021)
<sup>b</sup>Denotes chicken samples without breed information

Overall, we found a slightly lower prevalence of AMR genes (20% of sequenced isolates) than reports from the recent BYP-associated outbreaks led us to expect (8.5-42.9%; average of 28%) (CDC, 2021, 2022, 2023). It is possible that acquisition of AMR genes increases as the poultry mature and are exposed to environmental microbes carrying transmissible AMR genes.

This aligns with the increased frequency (41.4% of samples) of AMR genes reported in adult BYP (Patch et al., 2025). However, it also may simply reflect the serovar distribution in our sample sets, as even in adult BYP, we did not find AMR genes in *Salmonella* Enteritidis (Patch et al., 2025). Interestingly, none of our *Salmonella* Enteritidis isolates had the GyrA D87Y point mutation for ciprofloxacin/nalidixic acid, as this mutation was found in BYP outbreak-associated *Salmonella* Enteritidis isolates in 2022 (FDA, 2024a). However, only 93/1230 cases (7.5%) in 2022 had this point mutation (CDC, 2022; FDA, 2024a), so it is possible we simply did not have samples from the hatchery(s) carrying this mutation. Similarly, none of the *Salmonella* I 4,[5],12:i:-isolates we collected carried the *mphA* gene encoding azithromycin (FDA, 2024b); however, as this has not been reported in BYP samples, this is not surprising. Finally, despite numerous *Salmonella* Infantis isolates with plasmids, none of our isolates contained pESI-like plasmids carrying *bla*_CTX-M_ family genes, although it had been previously reported that ∼30% of *Salmonella* Infantis poultry isolates collected by FSIS and NARMS in 2021-2022 carried this gene (Li et al., 2024). This suggests that the plasmid may not have spread widely into the hatcheries supplying the BYP market.

Our study had several limitations. First, we were limited to stores that agreed to let us sample, and some had policies against participating in research. Second, we were limited by which hatcheries the stores purchased from; several major hatcheries were consequently not captured in this study. Third, we were limited by store practices. While we made strenuous efforts to communicate with stores and remind them to retain shipping boxes, some stores refused, preferring to provide the boxes to customers. In these instances, we sampled wood shavings, straw, cardboard, or newspaper in the tubs or stacked brooders the baby poultry were transferred to on arrival. These stores often combined breeds, particularly as baby poultry were sold. Even stores which typically complied occasionally forgot to retain boxes or keep the lids, which were labeled with breed information. These issues resulted in a high number of “unknown” samples. Finally, our SMP vs non-SMP analysis was limited by the fact that one major hatchery had multiple locations for the same breeds, and only some of the locations were enrolled in the SMP. Finally, drop shipping—or the practice of a hatchery filling orders from another hatchery, but shipping it under their own label—is known to occur, along with trans-shipping, or a hatchery obtaining birds from other hatcheries and mixing them prior to shipping; however, these practices are extremely difficult to track (Behravesh et al., 2014). Behravesh et al. noted that agricultural supply stores which order from one hatchery may actually receive birds from a mix of hatcheries without knowing it. We cannot rule out that drop or trans-shipping did not occur for the samples we collected; in fact, throughout the study, we would occasionally note labels from a different hatchery than the store normally purchased from when we sampled at that store.

Despite the limitations, our data indicate baby poultry frequently carry *S. enterica,* and they primarily carry clinically associated serovars, sometimes with concerning AMR genotypes. Effective communication of these risks is essential, particularly since outbreaks of live poultry-associated salmonellosis disproportionately affect children, partially because parents often view raising chickens as a good learning experience for children and may not be able to prevent children from cuddling animals or putting their hands in their mouths (Basler et al., 2016; Beam, Garber, Sakugawa, & Kopral, 2013; Elkhoraibi, Blatchford, Pitesky, & Mench, 2014; Larsen et al., 2022). Further research is needed on effective risk communication strategies for backyard poultry owners, agricultural/feed stores, hatcheries, and educators engaged in embryology projects. Ideally, this would coincide with changes to the SMP to increase *S. enterica* monitoring and implementation of *S. enterica* reduction strategies at hatcheries.

## Data Availability

Isolate data available upon reasonable request; sequences are posted on NCBI with accession numbers in Table S1.

---

**Table S1:** List of all sequenced *S. enterica* isolates including NCBI biosample numbers.

